# Three and Four Dimensional Characteristics of Color Doppler M-Mode Echocardiography

**DOI:** 10.1101/2023.11.07.23298198

**Authors:** Seyed Babak Moosavi Toomatari, Kamal Khademvatani, Seyedeh Zahra Karimi Sarabi

## Abstract

M-mode echocardiography is one of the basic modes of ultrasound imaging that is generally used in echocardiography. In this article we introduce a new approach to display three and four dimensional color M-mode echocardiography.

## Introduction

Edler and Hertz recorded heart movement by means of ultrasound in 1954 that displayed heart’s structures based on ultrasound waves and their reflections (1, 2, 5). But, two-dimensional echocardiography as a reliable modality to examine heart morphology and function was developed in 1970s and equipped by Doppler and also color Doppler in 1980s (3, 4).

Generally, B-Mode (brightness mode) and M-mode (motion mode) are two main modes of ultrasonography that display images of body structures. Unlike B-mode that is used in general imaging including heart imaging, M-mode is especially used in echocardiography (8). B-mode images are combination of pixels that each pixel has a position and brightness. The position is determined based on the depth of reflected echo and the brightness is determined based on the amplitude of reflected echo. So, real-time B-mode images are created by recording of each point gray-scale continuously (8, 9).

In M-mode echocardiography as an old echography mode, an ultrasonic wave is sent and received in a single line to produce one dimensional image. High frequency repetition of sending and receiving signal can produce a two dimensional image by placing a cursor on heart structures (3, 4).

When a sound source or receiver moves, frequency is changed; this change named as Doppler’s effect (8, 9, 10). Doppler effect is used to reveal the direction and velocity of blood flow (exactly blood cells). If blood cells move toward the transducer, the returning frequency will be higher than main frequency and vice versa (9).

Pulse wave (PW) Doppler, continuous wave (CW) Doppler, color Doppler and power Doppler are most common Doppler modes that used in medical purposes. One crystal is used to send and receive ultrasonic waves in pulse Doppler mode. PW cannot measure velocity of high blood flow. To solve this, CW Doppler mode is used that contains two crystals, one for sending and one for receiving ultrasound waves continuously (11, 12).

In medical utilization, direction and velocity of blood flow is displayed in colors: blue for blood flow direction away from transducer and red for blood flow toward transducer, conventionally (13, 14).

In this article we introduce a new approach to display and analyze color Doppler M-mode echocardiography of Descending aorta.

## Material and Method

We analyze color Doppler M-mode of descending aorta by using a vivid-6 echocardiography device. Color Doppler M-mode data is extracted at the beginning of descending aorta and saved as a two dimensional jpeg image in 24 bits RGB color format (Fig-1).

**Figure 1:**
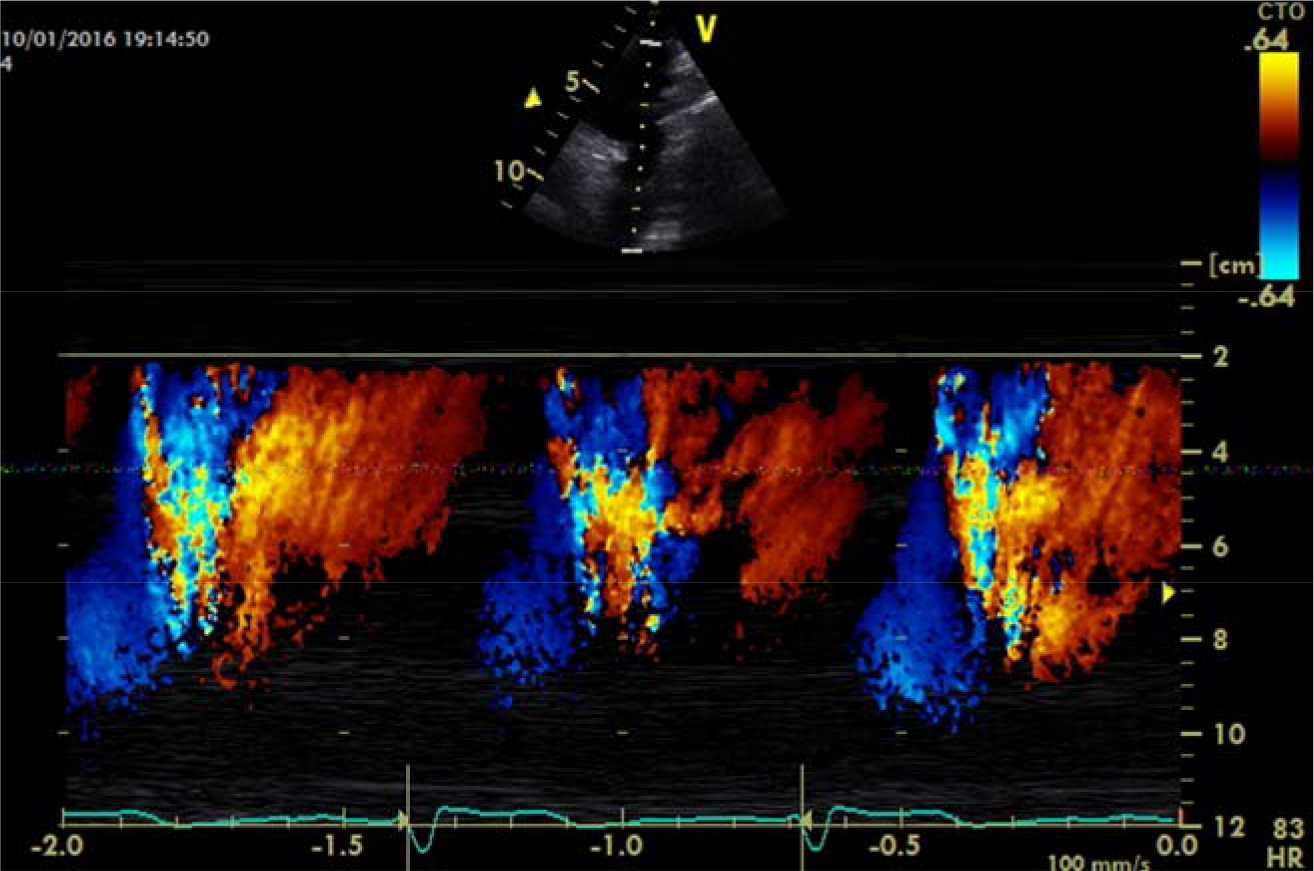
Color Doppler M-mode Echocardiography of Descending Aorta

We designed a Windows-based software to analyze doppler images. At the 1^st^ step, jpeg image is opened and one complete heart cycle is extracted in a rectangular that contains all Doppler data (Fig.2).

**Figure 2:**
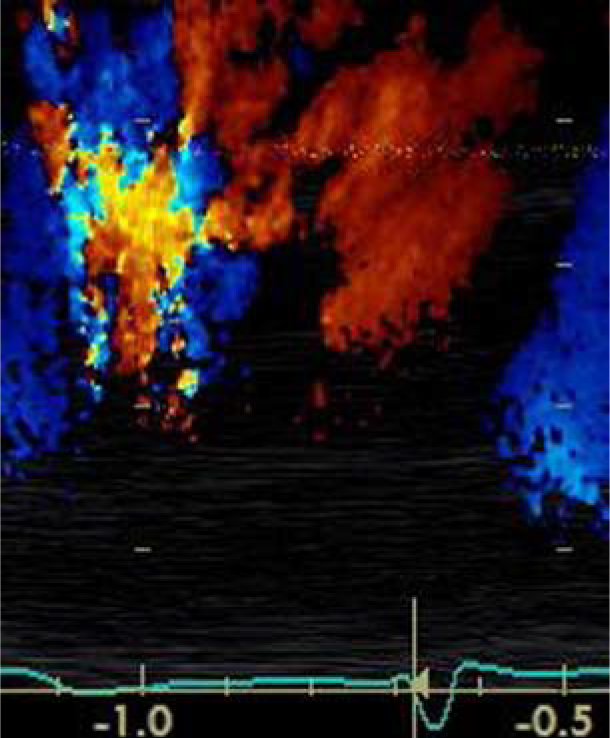
Extracted Data of One Complete Heart Cycle

At the 2^nd^ step extracted color-coded data of systole and diastole analyzed; each pixel which contains Doppler data is displayed on a three dimensional chart with X, Y and Z axis, where any axis divided into 256 equal parts (0 to 255). X axis represents green color, Y axis represents blue and Z axis represents red color of each pixel and a three dimension chart of color Doppler M-mode of descending aorta is displayed (Fig-3).

**Figure 3:**
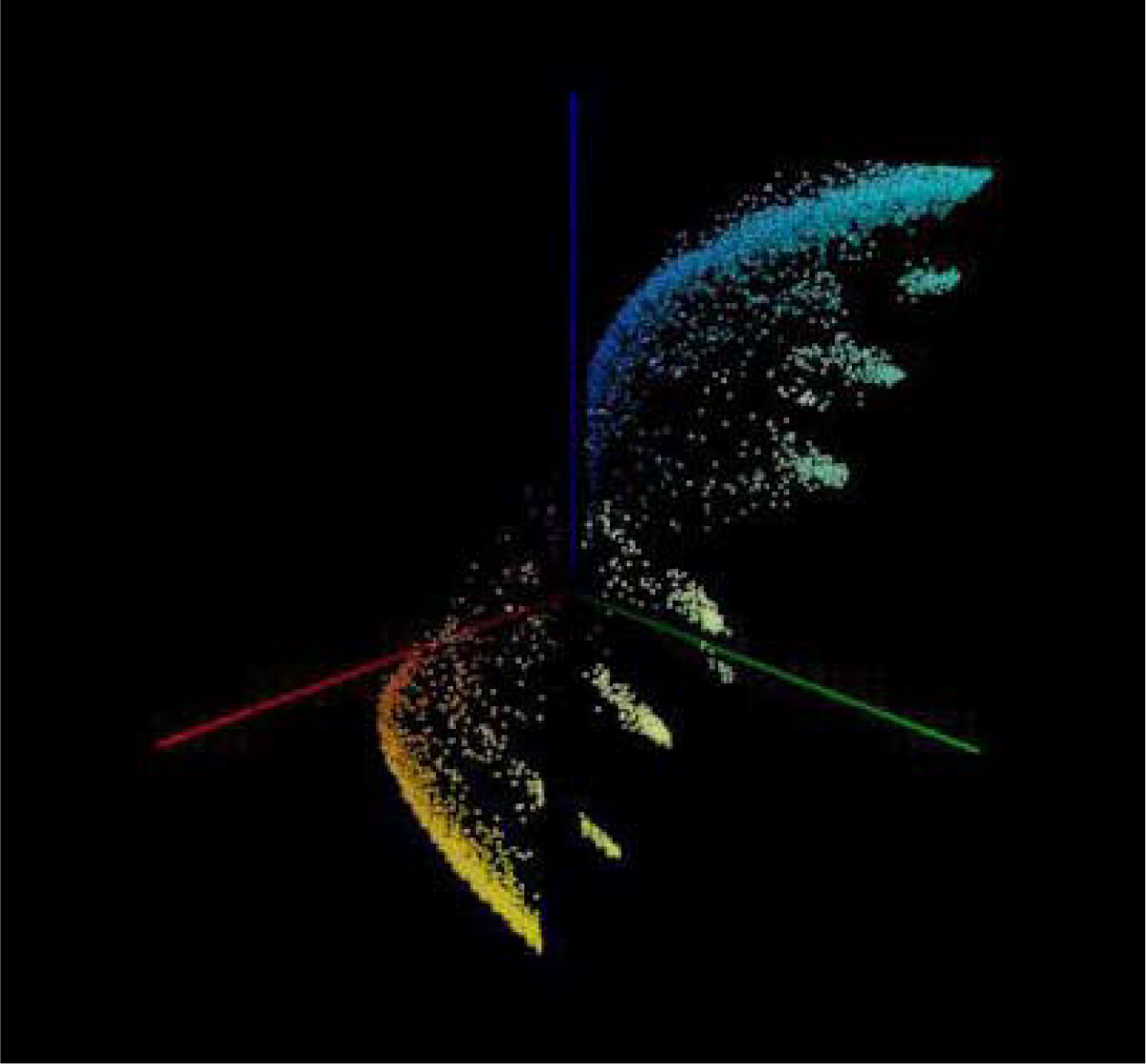
Three Dimensional View of Color Doppler M-mode Echocardiography

Now, we decide to display each pixel of color Doppler data in any time of a heart cycle. So, we spread a cycle of heart bit in 360 degree around Y axis. Therefore, 1 millisecond of a complete heart cycle will be equal to 360/duration. Finally, a four dimensional diagram will display as perspective, top and lateral view (Fig-4, Fig-5, Fig-6).

**Figure 4:**
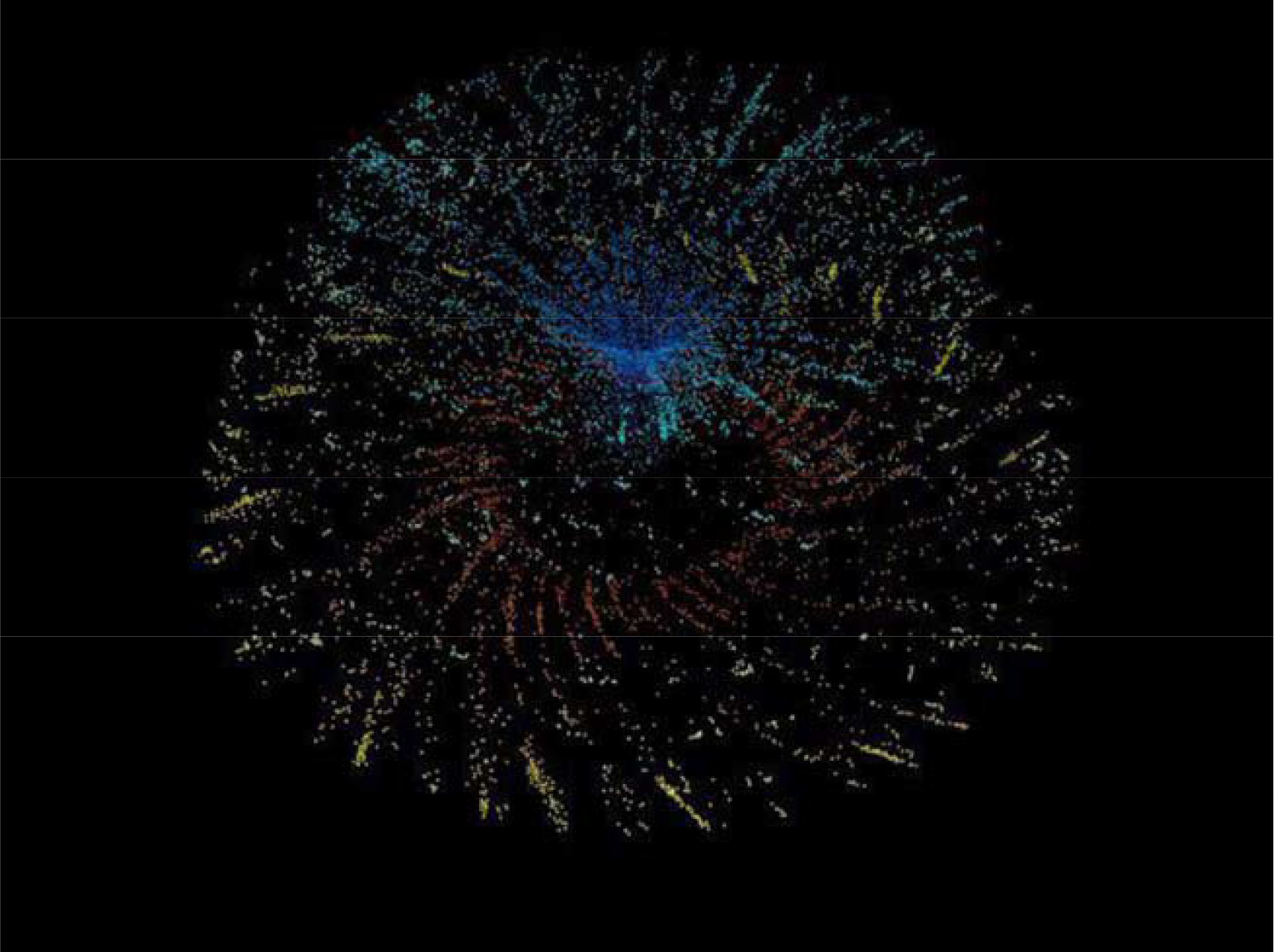
Four Dimensional View of Color Doppler M-mode Echocardiography, Perspective View

**Figure 5:**
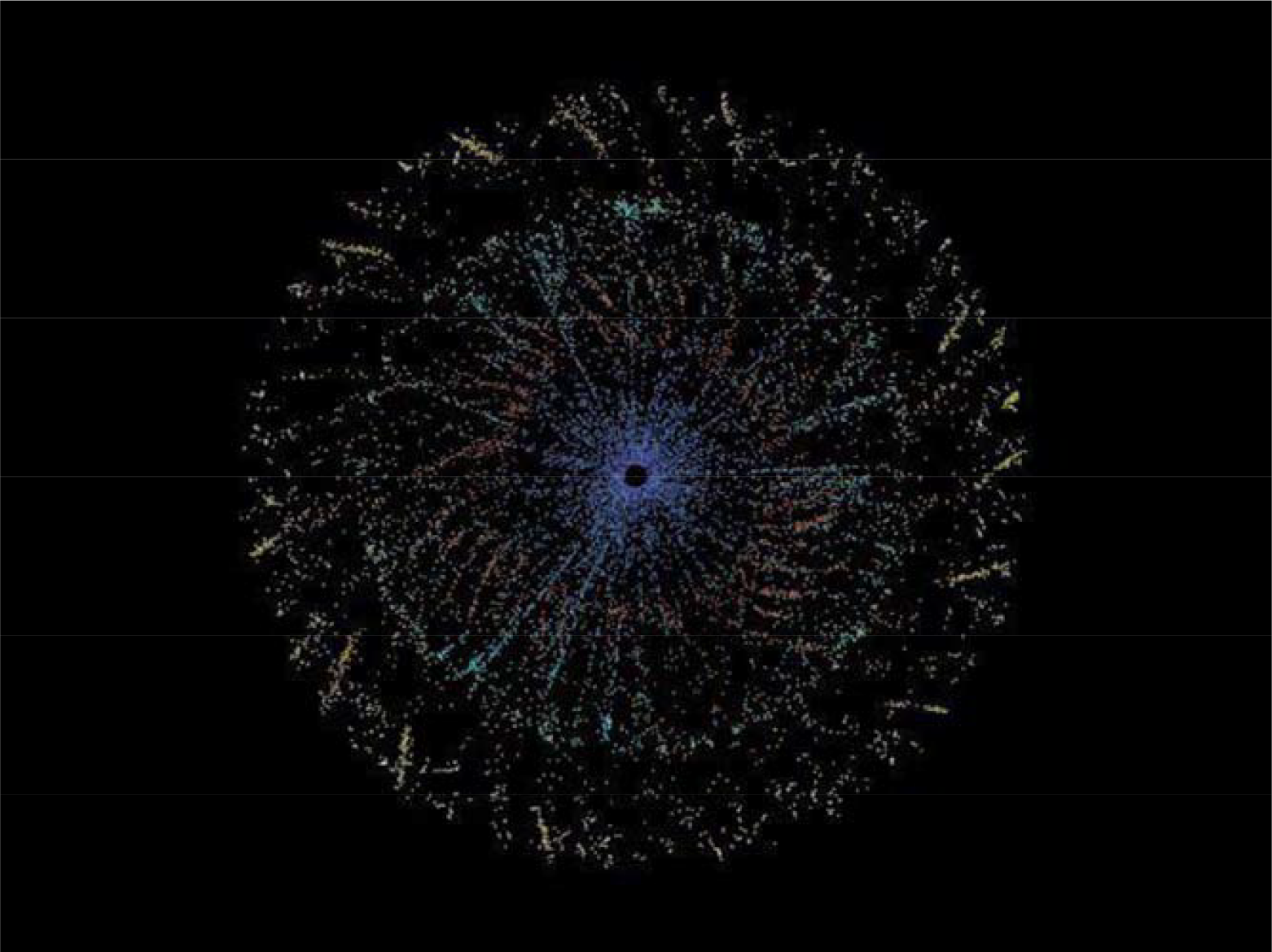
Four Dimensional View of Color Doppler M-mode Echocardiography, Top View

**Figure 6:**
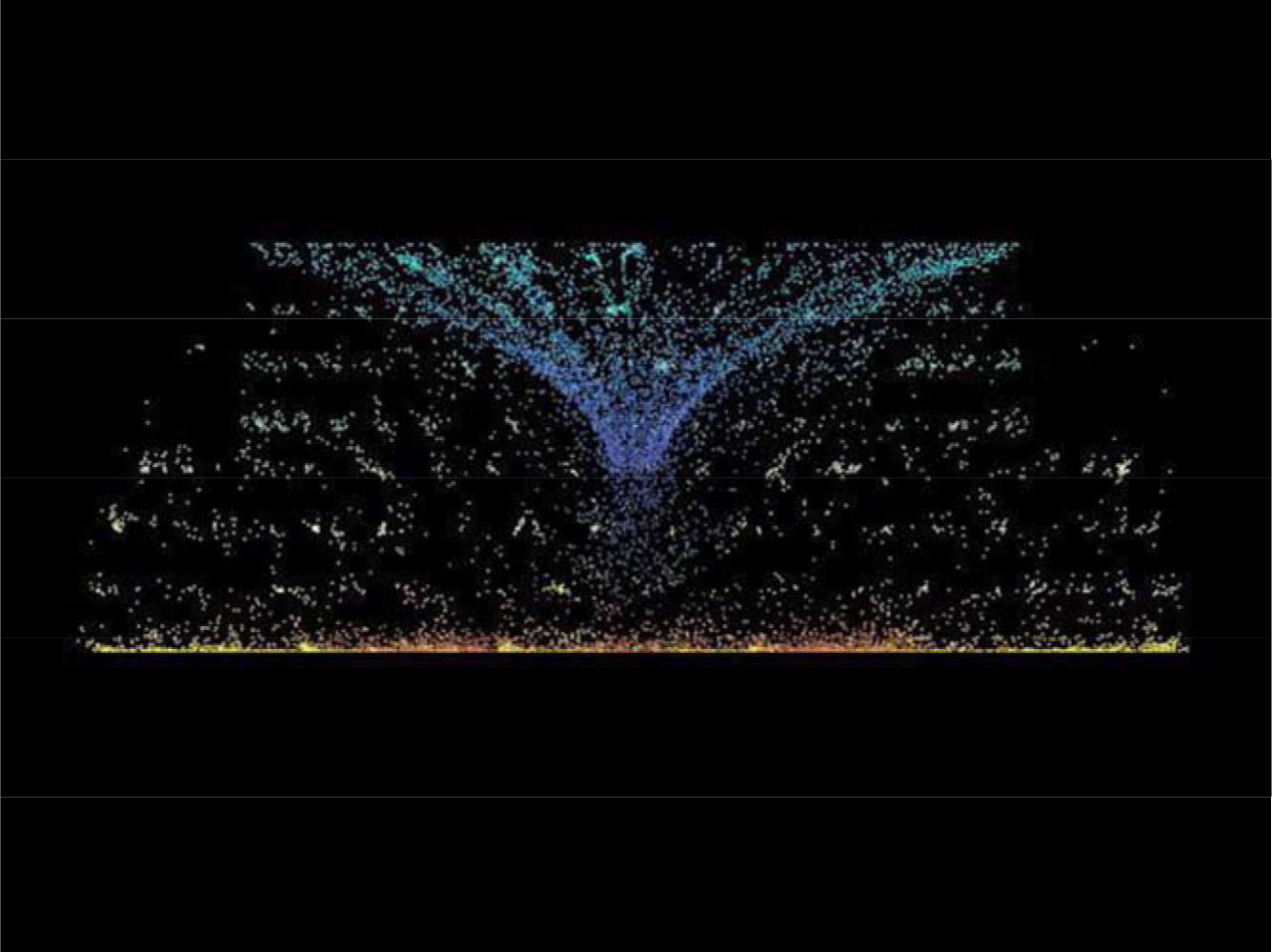
Four Dimensional View of Color Doppler M-mode Echocardiography, Lateral View

## Result

Two major flows (toward and away from transducer) and six minor flow (toward and away from transducer) is displayed in Fig-3. Major flows are main aortic flow and minor flows belong to first three branches of descending aorta. While turbulence of descending aorta is increased, such as aortic valve insufficiency, flow toward transducer is increased.

### Description

We introduce a new approach to display color Doppler M-mode echocardiography in three dimension and four dimension view. To quantified analysis at least four parameters could be extracted as number of pixels: systolic flow toward transducer (SFT), systolic flow away from transducer (SFA), diastolic flow toward transducer (DFT), diastolic flow away from transducer (DFA). We decide additional equation that may be useful: SFA/SFT, DFT/DFA, SFA/DFT and (SFA/SFT)/(DFT/DFA). Also, geometric analysis of color Doppler M-mode echocardiography such as sinus of angles α, β and δ may be valuable (Fig-7).

**Figure 7:**
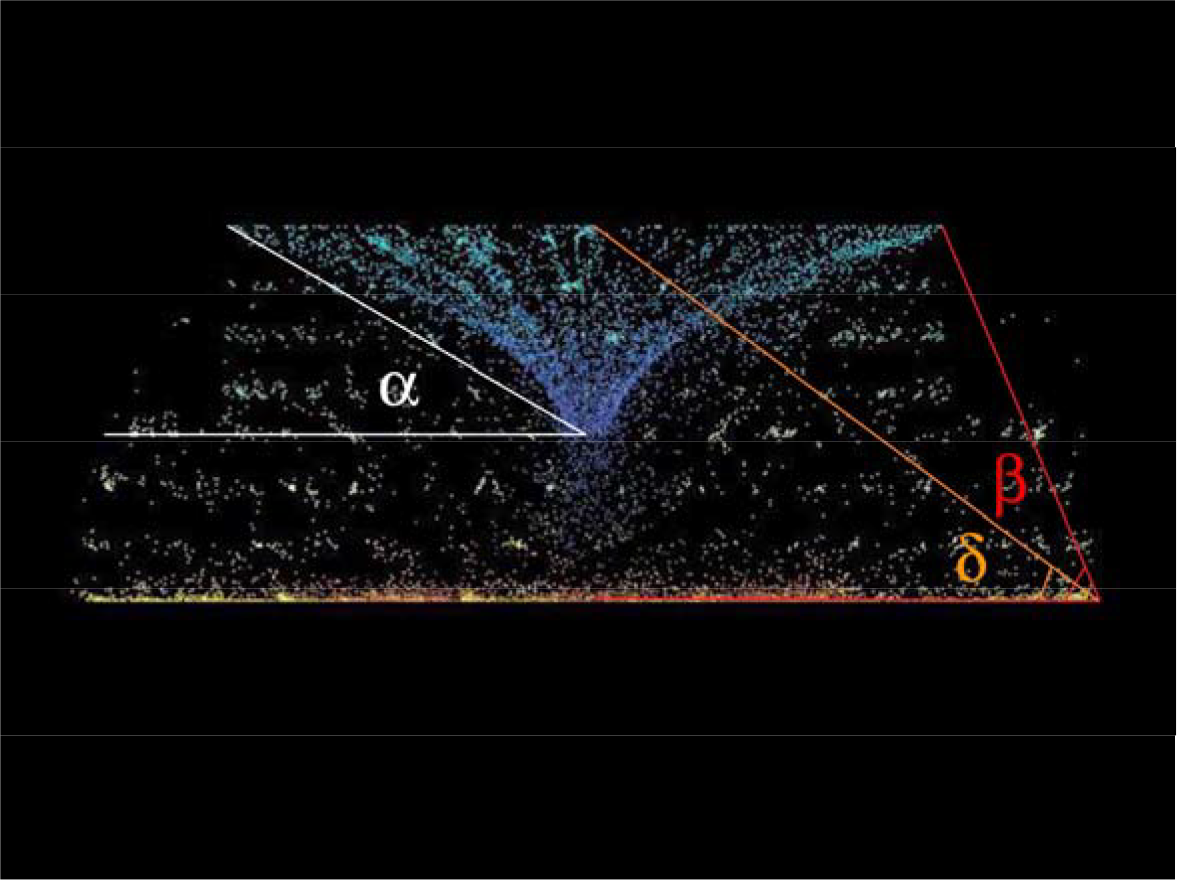
Geometric Data of color Doppler M-mode echocardiography

## Data Availability

All data produced in the present study are available upon reasonable request to the authors

